# Cross-Sectional Study of Periodontal Oral Dental Foci of Infection (PODFI) in Medically Compromised Patients at Lille University Hospital

**DOI:** 10.64898/2026.10.25.26366410

**Authors:** Solène Ribreux, Clara Roussel, Maxime Bedez, Éloïse Gilbert

## Abstract

Oral dental foci of infection (ODFI), particularly those of periodontal origin, can trigger systemic inflammatory responses that may cause bacteremia for medically compromised patients. Although guidelines recommend detection and elimination of these foci, precise data on the distribution of periodontal oral dental foci of infection (PODFI) risk within these populations remain scarce. This study aimed to describe periodontal sources of infection in medically compromised patients using a periodontal examination and follow-up form and a recently developed PODFI risk levels classification.

This observational, retrospective, cross-sectional, single-center study evaluated medically compromised patients with periodontitis in the dental surgery department of the Lille University Hospital between September 2024 and September 2025. Periodontitis was diagnosed according to the 2017 Classification of Periodontal Diseases established at the 2017 World Workshop. The primary outcome was the distribution of PODFI risk levels (moderate, high, very high). Secondary outcomes aimed to describe periodontal conditions in a specific population of medically compromised patients.

Thirty-nine patients were included in this study. Evaluation of PODFI risk (n = 34) revealed a predominance of moderate-risk profiles (67.6%) and a significant proportion of high-risk profiles (32.4%); no low-risk or very high-risk profiles were observed. Periodontal inflammation was very common (92%), with a median of 26 bleeding on probing (BOP) sites and 71 plaque-positive sites, showing a positive correlation (R = 0.52, p = 0.002).

Severe and active forms of periodontitis predominated (n = 39), with 74.4% of cases classified as stage III (28.2%) or stage IV (46.2%), and grade B was the most common (56.4%), followed by grade C (38.5%).

Medically compromised patients have a high prevalence of severe and active periodontitis, with nearly one-third of them classified as being at high risk for infection. The recently developed periodontal oral dental foci of infection (PODFI) classification provides a structured and clinically relevant tool for stratifying infection risk and guiding treatment decisions. Prospective multicenter studies are needed to confirm the validity of these PODFI risk levels.

## Introduction

Oral infections are a chronic source of inflammation and bacteremia that can affect systemic health [1]. In medically compromised patients, oral infections, particularly periodontal infections, can exacerbate or destabilize systemic conditions, especially diabetes [2] and cardiovascular diseases [3], and more than fifty other systemic conditions have been hypothesized to be related to periodontal diseases [4]. Periodontitis interacts with these systemic conditions by inducing or sustaining a systemic inflammatory response linked to local inflammation, the dissemination or translocation of pathogens, and cross-reactivity [5,6]. Furthermore, the literature suggests that periodontitis is associated with an increased risk of all-cause mortality, with affected patients exhibiting more than a twofold higher risk of death (HR = 2.19) [7], highlighting its clinical relevance in medically compromised patients.

In this context, the identification and management of periodontal oral dental foci of infection (PODFI) appear essential for improving systemic health. In France, the French Society of Oral Surgery (SFCO) defines an oral dental focus of infection (ODFI) as the presence of an oral bacterial focus, regardless of the presence or absence of clinical manifestations at the time of evaluation. These ODFIs include, in particular, endodontic and periodontal infections [8]. SFCO therefore recommends screening and eliminating these foci in medically compromised patients.

To optimize this approach, an initial study led to the development of a standardized periodontal examination and follow-up form, incorporating concepts from the 2017 Classification of periodontal diseases and a classification of PODFI. This periodontal form allows both for determining the periodontal diagnosis, classifying the patient’s PODFI risk ranging from low to very high, and for tailoring the patient’s clinical pathway based on the urgency of their medical condition and the feasibility of plaque control. The form can also be used for the follow-up.

Little data is available regarding the distribution of PODFI risk levels among medically compromised patients, whether based on older or more recent classifications of periodontal diseases. However, a better characterization of these infection foci appears necessary to adapt treatment strategies and prioritize care based on the patient’s medical context.

The aim of this study was to describe and quantify PODFI in medically compromised patients with periodontitis. The primary objective was to assess the distribution of PODFI risk levels according to the proposed classification. The secondary objectives were to characterize periodontal conditions in a population of medically compromised patients.

## Materials and Methods

This single-center retrospective cross-sectional observational study was conducted in the specialized medically compromised patients periodontal care service of Dental Service of hospital-based dental school of Lille University Hospital.

The study population was patients with suspected periodontal disease who attended their first consultation at this specialized care service between September 1, 2024, and September 27, 2025, and identified as medically compromised patients.

Only adults (≥ 18 years) presenting a high-risk medical condition warranting care within the Department of Dentistry at the Lille University Hospital, with a confirmed diagnosis of periodontitis were included.

Patients who refused to allow their medical data to be used were excluded.

Data were collected retrospectively from patients’ paper standardized periodontal examination form. Information necessary for the study was extracted and recorded in a pseudonymized database. In cases of missing data, these were excluded from the corresponding statistical analyses.

This study was conducted in accordance with regulations regarding the protection of personal health data. In fact, the study protocol was submitted to the Data Protection Officer (DPO) of the Lille University Hospital and received a favorable opinion from the institution’s ethics committee (reference: 2025-930-S140). An information letter was also made available to patients on the Lille University Hospital website to allow them to exercise their right to object to the use of their data if they wished.

The variables collected included the diagnosis of periodontitis, the PODFI risk level, the presence of periodontal inflammation and suppuration, the number of sites with bleeding on probing, the number of plaque-positive sites, the presence of endo-periodontal lesion, the periodontal diagnosis, the number of missing teeth, and the type of treatment pathway assigned to the patient.

The primary outcome measure was the PODFI risk level, determined according to the aforementioned classification.

Secondary evaluation criteria included the presence of periodontal inflammation and suppuration, the number of sites with bleeding on probing, the number of plaque-positive sites, the presence of endo-periodontal lesion, the periodontal diagnosis (stage and grade), the number of missing teeth, and the type of treatment pathway.

The diagnosis of periodontitis was made in accordance with the classification established at the 2017 World Workshop in Chicago. It is based on the presence of interdental clinical attachment loss in at least two non-adjacent teeth or clinical attachment loss ≥ 3 mm in the buccal, palatal, or lingual regions associated with a pocket depth > 3 mm on at least two teeth. This clinical attachment loss cannot be attributed to any cause other than periodontitis [9]. This classification is based on a staging system (stages I to IV) describing the severity of the disease and the complexity of management, and a grading system (grades A to C) indicating the progression and risk of progression of periodontal disease, incorporating, among other factors, grade-modifying risk factors such as smoking or diabetes [9].

The PODFI risk level was determined for each patient based on the previously proposed classification for the Evaluation of Periodontal Oral Dental Foci of Infection (C. Roussel *et al.*, manuscript in preparation) and detailed in the table below. As only patients with periodontitis were included, the low risk could only be represented by stabilized periodontitis.

Sample size calculation was based on the estimated prevalence of very high-risk periodontitis. Assuming that 23% of patients with periodontitis would be classified as very high risk [10], a minimum sample size of 69 patients was required to estimate this proportion with a 95% confidence level and a 10% margin of error in a reference population of approximately 3 million medically compromised patients with periodontitis. This estimate was derived from French national health insurance data reporting approximately 7 million individuals with a long-term condition [11] combined with an estimated periodontitis prevalence of 44% in the general population [10].

Missing data were excluded from the analyses. Categorical variables were described using frequencies and percentages. Quantitative variables were summarized as mean ± standard deviation (SD) when normally distributed, or as median and interquartile range (IQR) otherwise. Data distribution was assessed using the Shapiro–Wilk test. Correlation analyses were performed using statistical methods appropriate to the distribution of the data: Pearson’s correlation coefficient was used when both variables were normally distributed, whereas Spearman’s rank correlation coefficient was used when at least one variable did not follow a normal distribution. A chi-square goodness of fit test was used to assess whether the observed prevalence across categories differed significantly from an expected uniform distribution, assuming equal proportions (33.3%) in each category. Statistical analyses and graphical representations were performed using Microsoft Excel 365 (Microsoft Corp., Redmond, WA, USA) and GraphPad Prism (11.0.2; GraphPad Software, San Diego, USA). The significance threshold was set at p < 0.05.

## Results

Forty-one records were initially collected. Two records were excluded due to an incomplete periodontal examination that did not allow for the determination of the PODFI risk level, as the “charting” section of the form had not been completed. In total, 39 records, corresponding to 39 patients, were included. Analyses were performed on the available data; sample sizes therefore vary between variables due to missing data.

The results are presented by distinguishing the PODFI risk level, corresponding to the primary endpoint, followed by the various associated periodontal parameters.

Among the 34 patients for whom risk-level data were available, 23 (67.6%) were classified as moderate risk and 11 (32.4%) as high risk, while no patients were classified as low or very high risk (Table 3). The chi-square goodness of fit test, with an expected uniform distribution, indicated that the observed prevalence differed significantly from the expected distribution (p < 0.0001).

**Table 1:** PODFI Risk Level.

| PODFI Risk Level | Diagnostic Criteria |
| --- | --- |
| Low | Gingivitis or stabilized periodontitis |
| Moderate | Active periodontitis with:<br>< 100 sites with BOP<br>AND<br>< 100 plaque-positive sites |
| High | Active periodontitis with:<br>≥ 100 sites with BOP<br>AND / OR<br>≥ 100 plaque-positive sites |
| Very High | Active periodontitis with:<br>Suppuration<br>AND / OR<br>Endo-periodontal lesion |

**Table 2:**
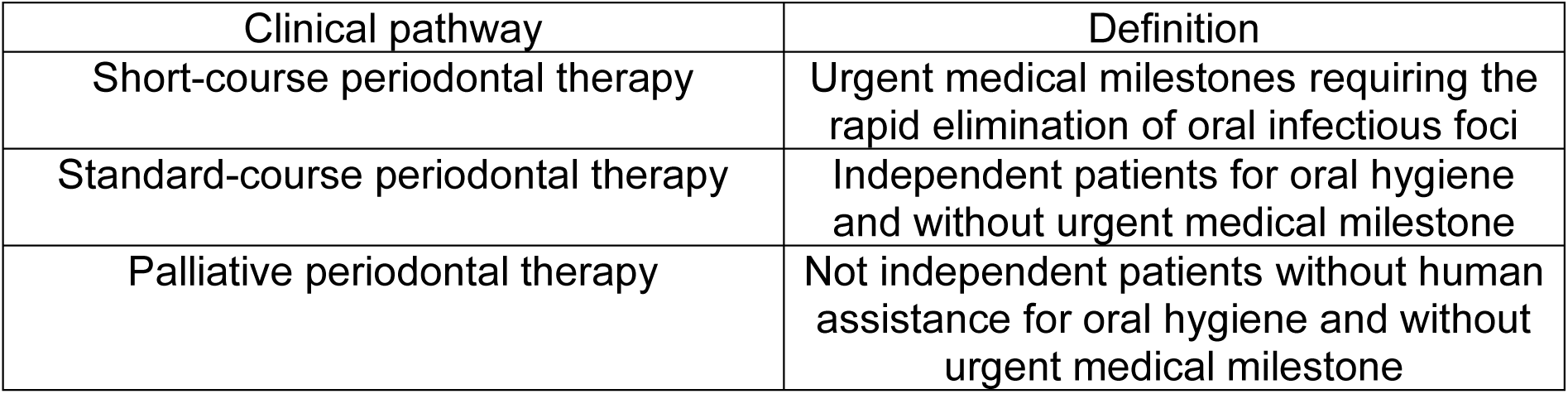
Type of clinical pathway.

| Clinical pathway | Definition |
| --- | --- |
| Short-course periodontal therapy | Urgent medical milestones requiring the rapid elimination of oral infectious foci |
| Standard-course periodontal therapy | Independent patients for oral hygiene and without urgent medical milestone |
| Palliative periodontal therapy | Not independent patients without human assistance for oral hygiene and without urgent medical milestone |

**Table 3:** Patients’ PODFI levels (moderate: <100 bleeding sites and plaque-positive sites; high: ≥100 bleeding sites and/or plaque-positive sites; very high: presence of suppuration and/or endo-periodontal lesions)

| PODFI level | Number of patients | Percentage |
| --- | --- | --- |
| Low | 0 | 0 |
| Moderate | 23 | 68 |
| High | 11 | 32 |
| Very high | 0 | 0 |

Periodontal inflammation was observed in 92% of patients for whom this data was available (n = 37). The median number of sites showing bleeding on probing was 26 [16; 73] (n = 39), while the median number of plaque-positive sites was 71 [40; 122] (n = 34). The number of sites showing bleeding on probing and plaque-positive sites varied between patients with ranges from 4 to 157 and from 0 to 186 respectively. No cases of suppuration or endo-periodontal lesions were observed during the periodontal examination.

A positive correlation between the number of sites showing bleeding on probing and the number of plaque-positive sites was observed using Pearson’s correlation coefficient (R = 0.52; p = 0.002; n = 33 complete pairs, due to one missing value) (Table 4 and Table 5), also illustrated in Figure 1.

**Figure 1:**
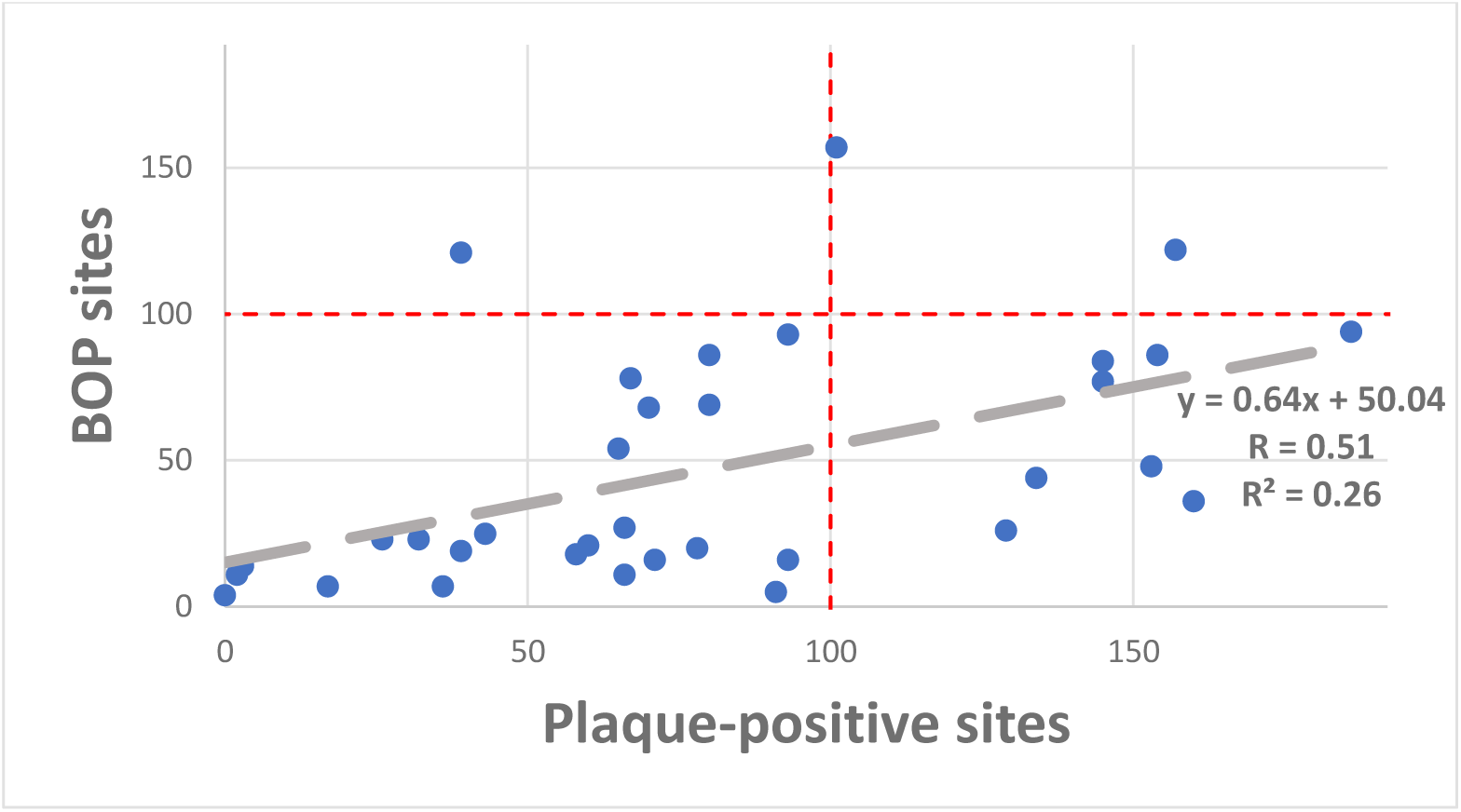
Correlation between the number of sites with bleeding on probing and the number of plaque-positive sites.

**Table 4:**
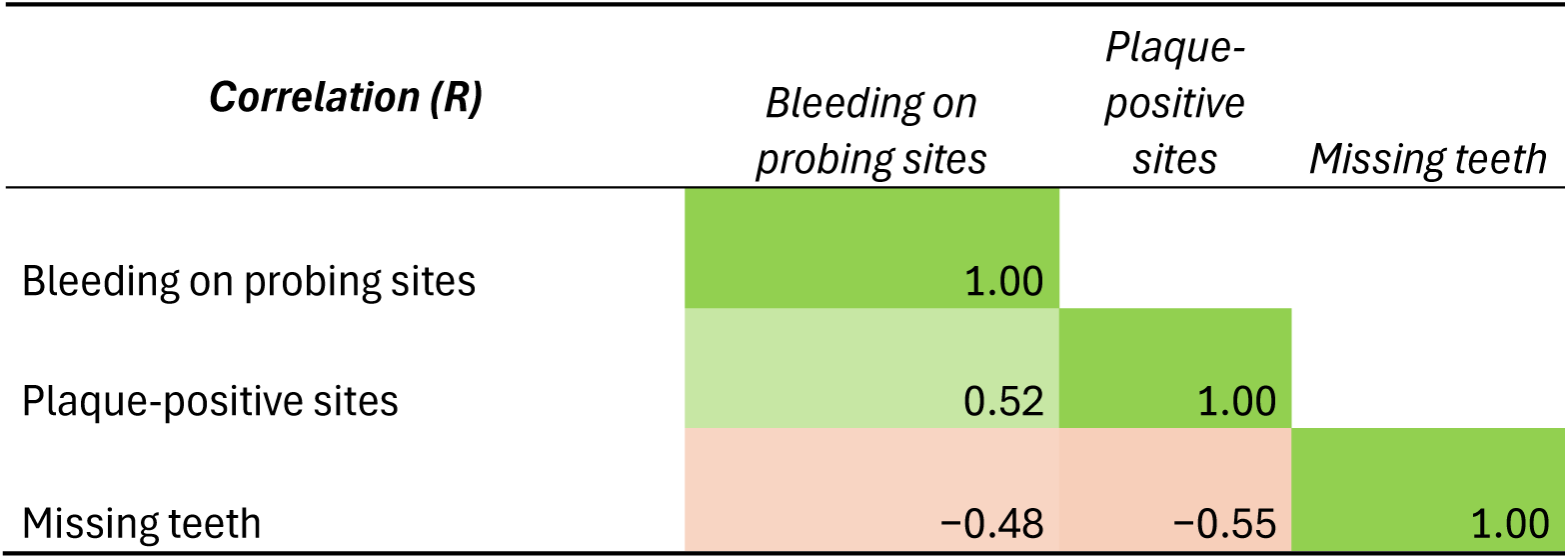
Relationship between the number of sites with bleeding on probing, the number of plaque-positive sites and the number of missing teeth.

**Table 5:** Relationship between the number of sites with bleeding on probing, the number of plaque-positive sites and the number of missing teeth with correlations.

|  | **R** | **R<sup>2</sup>** | **p** | **Number of valid pairs** |
| --- | --- | --- | --- | --- |
| **BOP vs. Plaque** | 0.52 | 0.27 | 0.002 | 33 |
| **Missing teeth vs. BOP** | -0.48 | 0.23 | 0.002 | 39 |
| **Missing teeth vs. Plaque** | -0.55 | 0.30 | 0.001 | 33 |

The median number of missing teeth was 12 [6; 18]. A significant negative correlation was demonstrated, using Pearson’s correlation coefficient, between the number of missing teeth and the bleeding index (R = −0.48; p = 0.002), as well as between the number of missing teeth and the number of plaque-positive sites (R = −0.55; p = 0.001) (Table 4).

Figure 2 shows that the study population consisted mostly of patients with severe periodontitis (n = 29; 74.4%), with a predominance of stage IV (n = 18; 46.2%) and stage III (n = 11; 28.2%), while stage II (n = 8; 20.5%) and stage I (n = 2; 5.1%) were less common. Regarding grade, grade B was the most common (n = 22; 56.4%), followed by grade C (n = 15; 38.5%). In particular, stage IV was represented exclusively by grades B (n = 8) and C (n = 10).

**Figure 2:**
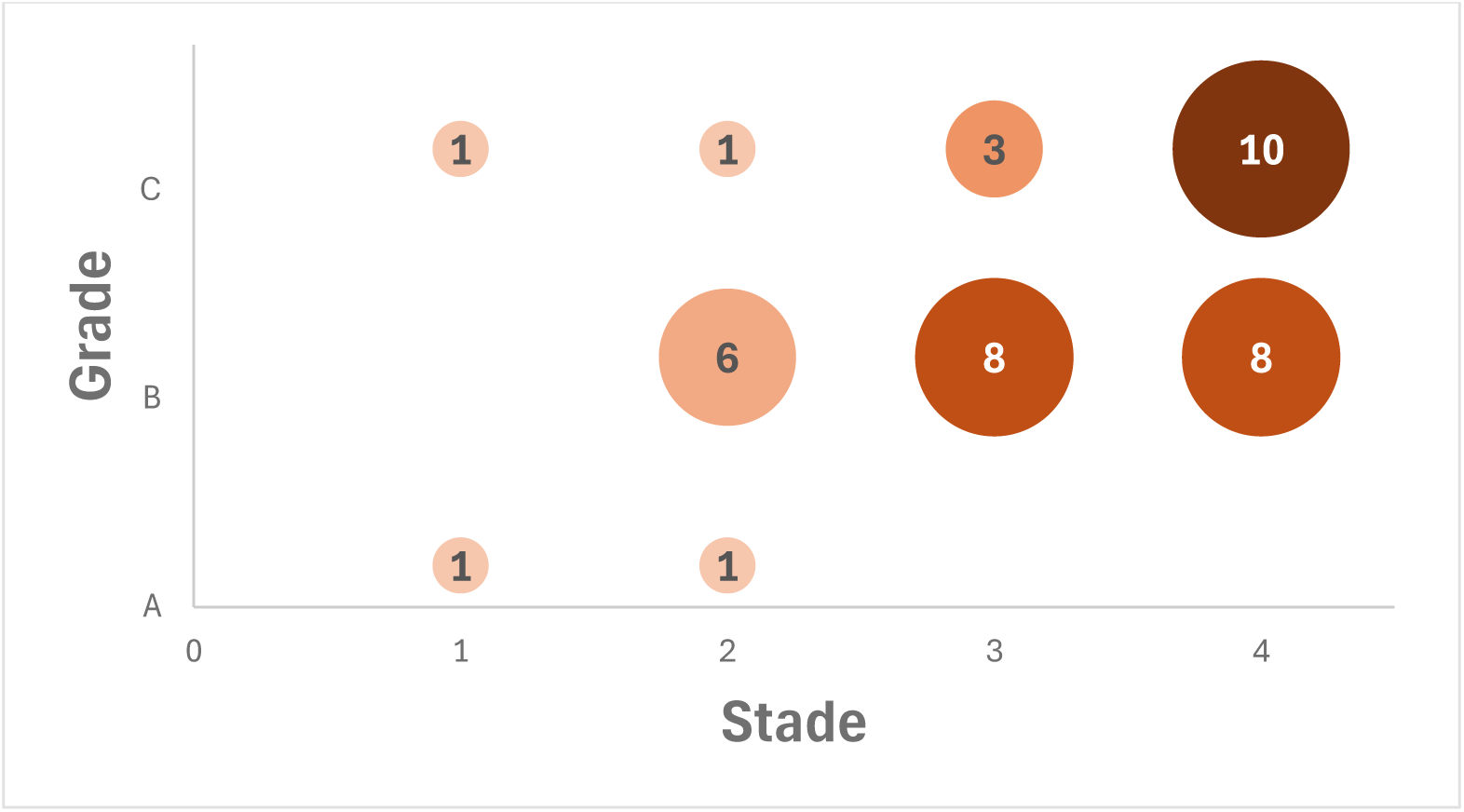
Distribution of patients by stage and grade of periodontitis according to the 2017 Chicago World Workshop classification.

Finally, the distribution of patients by clinical pathway showed a slightly higher frequency of standard-course (n = 16; 51.6%), followed by short-courses (n = 14; 45.2%), while palliative periodontal therapy courses (patients not independent for oral hygiene [OH]) were underrepresented (n = 1; 3.2%). The joint analysis of the number of sites showing bleeding on probing and the number of plaque-positive sites by clinical pathway is presented in Figure 3. The threshold lines defined along the X and Y axes allow for the distinction between moderate-risk profiles, located below both thresholds, and high-risk profiles where one of the values exceeds one of the thresholds. Most patients in the short pathway (n = 11; 78.6%) were in the moderate-risk zone, with low to intermediate values for both parameters. However, two patients in the short-term clinical pathway had high biofilm values, placing them in the high-risk zone, and the risk level could not be determined for one patient owing to missing plaque-positive site data. Interpretation of the results regarding patients in the palliative clinical pathway remains limited due to the small sample size in this group.

**Figure 3:**
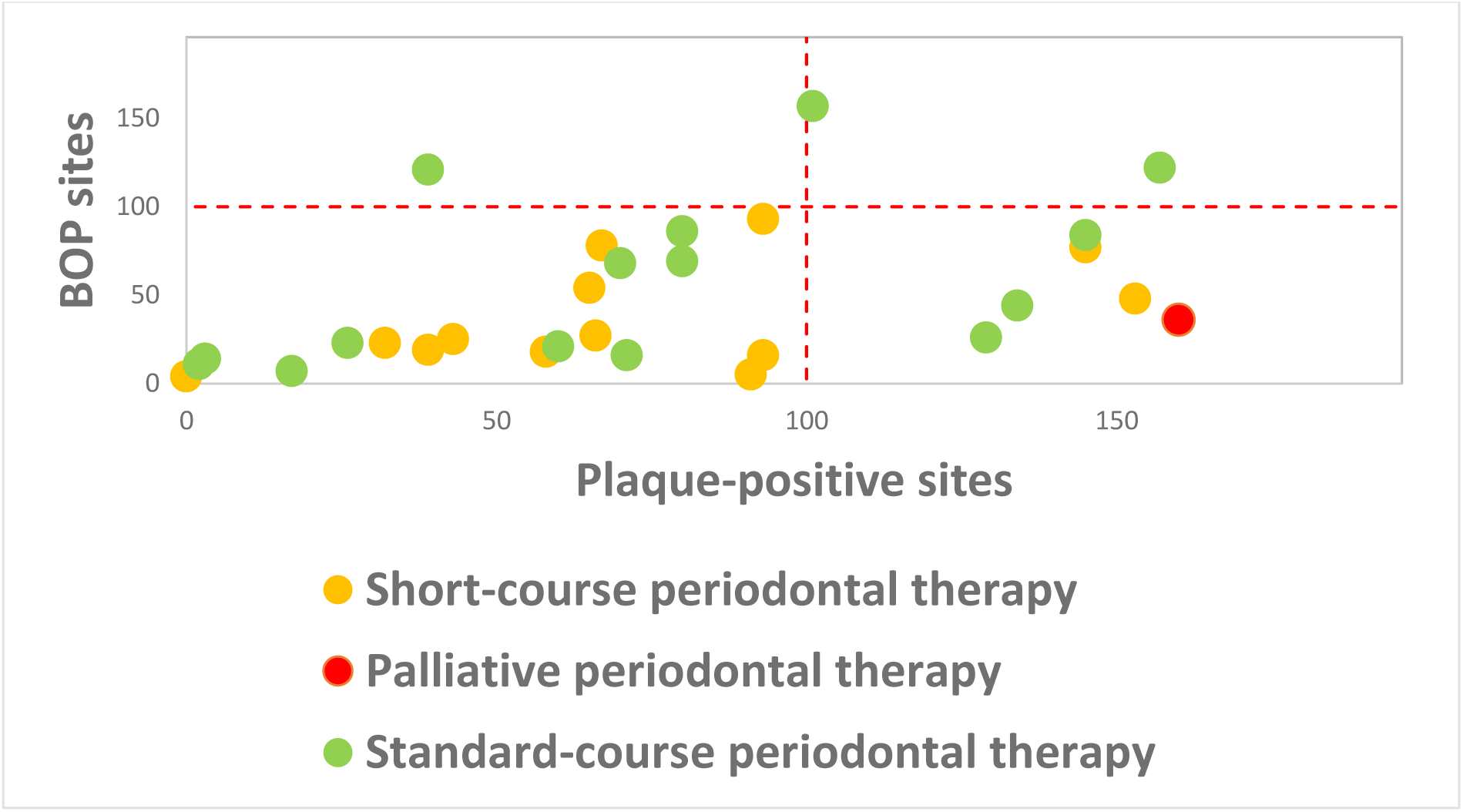
Joint analysis of the number of sites showing bleeding on probing and the number of plaque-positive sites, by clinical pathway.

## Discussion

The objective of this study was to assess the severity of periodontal infection foci in medically compromised patients using a new periodontal examination and follow-up form.

Classification of patients by risk-level showed a predominance of moderate-risk profiles, with a notable proportion of high-risk patients (32.4%). The chi-square goodness of fit test indicates that the distribution of individuals across categories was not uniform. The absence of low-risk patients was consistent with the study’s inclusion criteria which excluded gingivitis, and stabilized periodontitis was not expected as patients came for periodontal care. This distribution was also consistent with the median values observed in the population, with 26 [16; 73] sites showing bleeding on probing and 71 [40; 122] sites showing biofilm, corresponding to a moderate level of infection according to the proposed classification (C. Roussel *et al.*, manuscript in preparation). Despite this predominance of moderate-risk profiles, nearly one-third of patients were at high risk, highlighting the persistence of significant PODFI within this medically at-risk population. By examining the distribution of patient clinical pathways according to risk level, it was shown that the only patient in a palliative clinical pathway, a non-independent patient without human assistance for oral hygiene and without urgent medical milestone, had a high level of biofilm, with 160 sites exhibiting biofilm, which classified him into the high-risk category. These results fit into a broader context of loss of autonomy regarding oral hygiene. In France, approximately 1.3 million elderly people are in a state of dependency [12]. Among dependent elderly people living in care facilities, it has been shown that 60 to 70% cannot perform their oral hygiene independently, while another study reports that 27% of residents require partial or total assistance with hygiene [13]. However, dependence on others for oral hygiene is not limited to the elderly. International data on patients with disabilities have shown that 40.1% of patients require family assistance with brushing and 8.8% depend on an external caregiver [14]. This highlights the need for human assistance for patients with motor deficits. However, the interpretation of results regarding patients on long-term clinical pathways without OH remains limited due to the small sample size in this group.

Among patients in the short-term clinical pathway, who face urgent medical milestones (e.g., a hematopoietic stem cell transplant), the proportion of patients classified as high-risk was solely due to having more than 100 sites with biofilm. This data suggests that education on effective plaque control could help reduce their risk of oral and periodontal infections. This hypothesis is supported by the work of Lockhart et al., who reported that a plaque score ≥ 2 increased the risk of post-brushing bacteremia by a factor of 3.78, while a calculus score ≥ 2 increased this risk by a factor of 4.43 [15]. These data confirm the importance of biofilm control in reducing the risk of systemic infection.

The results revealed a high prevalence of severe and active periodontitis within the study population. The majority of patients presented with advanced stages (III and IV), associated with grades B and C, indicating a disease that is both extensive and progressive. This observation was consistent with the profile of the included patients, characterized by the presence of systemic conditions that could interact with periodontal disease.

The presence of inflammation in 92% of patients is consistent with the active nature of the periodontitis observed in this study. Indeed, the classification resulting from the 2017 World Workshop defines periodontitis as a chronic inflammatory disease [9], with gingival inflammation serving as a marker of disease activity. Furthermore, Van Dyke and Kornman emphasize the central role of the inflammatory response in periodontal pathophysiology. According to them, disease progression depends not only on bacterial presence but also on the intensity and regulation of the immuno-inflammatory response [16]. Thus, the high prevalence of inflammation observed in this population of medically at-risk patients suggested persistent inflammatory activity that could contribute to the maintenance of periodontal infection foci.

The high prevalence of inflammation and the high number of sites showing bleeding on probing confirmed disease activity. The median values for the number of sites showing bleeding on probing and the number of sites showing biofilm confirmed inflammatory activity associated with a high bacterial load within the study population. The positive correlation observed between these two parameters (R = 0.52; p = 0.002) supported the hypothesis of a close link between bacterial load and inflammatory activity, consistent with current pathophysiological data [15,17,18]. However, this correlation remained moderate, suggesting the involvement of other factors in the clinical manifestation of the disease.

For example, the probing bleeding index may be altered by tobacco use. Smoking has been associated with an altered gingival vascular response. Indeed, it causes vasoconstriction and impaired gingival circulation, leading to a decrease in local blood flow [19]. A cohort study demonstrated the impact of this mechanism, showing that heavy smokers had 44% lower odds of bleeding on probing at shallow sites (0–3 mm) than non-smokers (OR = 0.56), potentially masking underlying periodontal inflammation [20]. Furthermore, smoking has been associated with an impaired immune response, with increased levels of neutrophil enzymes including myeloperoxidase, β-glucuronidase and neutrophil elastase, reflecting an over-activated host response [21]. It has also been shown to disrupt the gingival epithelial barrier by altering the expression of junctional adhesion molecule-1 (JAM-1), increasing epithelial permeability to bacterial components such as LPS and peptidoglycan [22].

In contrast, no suppuration or endo-periodontal lesions were observed in the study population. This finding, combined with the absence of very high-risk profiles, suggested that most patients had active chronic forms of the disease rather than acute or exacerbated forms. Indeed, endo-periodontal lesions remain rare: while a study based on the 2017 classification reports a prevalence of approximately 4.9% [23], another study using CBCT imaging on 800 patients with periodontitis, most of whom were at stage III, shows slightly higher values, reaching 9.62% [24]. This distinction between chronic and acute manifestations of periodontal disease was important in the context of managing PODFI, as chronic forms may be less clinically apparent but nonetheless systemically harmful. Similarly, suppuration on probing is a relatively rare and localized sign in periodontitis. A 2013 study reported a prevalence of 24.4% of suppuration observed in patients with chronic periodontitis and 30.3% in what were then termed aggressive forms, whereas among these patients, on average, only 5.72% and 6.96% of probed sites exhibited suppuration, showing suppuration as a local phenomenon [25]. However, these acute forms, although less common, are generally associated with a high bacterial load and an increased potential for systemic spread, underscoring the importance of identifying them in at-risk populations [26].

The bacterial load may also vary depending on the amount of dental plaque. The O’Leary plaque index, initially based on the assessment of four tooth surfaces (mesial, buccal, distal, lingual) [27], was adapted in this study to six sites per tooth (mesio-buccal, buccal, disto-buccal, disto-lingual, lingual, mesio-lingual) to increase the sensitivity and accuracy of the measurement. Indeed, contemporary periodontal approaches rely on a six-site analysis [28], allowing for better detection of local variations in biofilm, particularly at the interproximal level, and limiting the risk of underestimation associated with simplified indices. This adaptation is part of a methodological approach aimed at improving the reproducibility and validity of measurements in a clinical research context. The number of plaque-positive sites highlighted heterogeneity in plaque control within the population, with a minimum of none to a maximum of 186 sites.

Including the number of missing teeth provided additional insight into the history of the disease. The negative correlations observed between the number of missing teeth and plaque and bleeding parameters did not indicate improved periodontal health but rather reflected the reduced number of remaining teeth and examinable sites available for clinical assessment. Thus, one might hastily conclude that reducing the number of remaining teeth could lower the PODFI risk. However, it must not be forgotten that a decrease in the masticatory index is correlated with increased mortality; in the study by Peng *et al.*, each loss of 10 teeth was associated with a 15% increase in the risk of all-cause mortality [29]. Complete edentulism increases the risk of all-cause mortality by 66% [7]. Indeed, patients who must adapt their diet due to reduced masticatory capacity may sometimes no longer be able to obtain dietary intake that meets their nutritional needs. A 2022 meta-analysis showed that non-functional dentition, particularly having fewer than 20 teeth, was associated with a 21% increased risk of malnutrition in older adults (OR = 1.21) [30]. These data underscore the importance of preserving functional dentition as part of a comprehensive approach to health.

However, several limitations of this study should be noted. The single-center design and small sample size limited the generalizability of the results. In particular, only 39 patients could be included, whereas the sample size calculation required 69, and no very high-risk profile was observed although a prevalence of 23% had been assumed; the study was therefore underpowered to estimate this proportion precisely. Furthermore, the use of retrospective data from paper medical records posed a risk of information bias.

Additionally, the exclusion of missing data in the analyses could constitute a source of bias, particularly if these data were not randomly distributed. This approach could lead to altered estimates and limit the representativeness of the results.

The use of absolute values for certain parameters, particularly the number of sites with biofilm and bleeding, could be considered a methodological limitation. Since these measures are directly dependent on the number of teeth present, they were influenced by the number of examinable sites, introducing a measurement bias. However, from the perspective of assessing infection risk, the use of the absolute number of affected sites appeared relevant. Indeed, bleeding on probing is a site-specific parameter reflecting local inflammation associated with bacterial activity [31], with each affected site corresponding to an area of epithelial ulceration capable of facilitating bacterial translocation into the bloodstream [32]. Thus, an increase in the number of affected sites indicates an increase in the inflammatory burden and the surface area capable of allowing bacterial translocation, regardless of the total number of teeth [33]. Conversely, expressing the data as a percentage may lead to an underestimation of the actual risk [34].

Furthermore, this study did not allow for the direct assessment of bacteremia or the identification of the bacterial families involved in these bacterial translocations. The absence of microbiological analyses thus constitutes a significant limitation in establishing a direct link between the observed clinical parameters and the actual systemic infection risk. Indeed, Forner *et al*. demonstrated that transient bacteremia could occur after toothbrushing, chewing, and scaling in patients with periodontal inflammation. The authors demonstrated that the frequency of bacteremia increased with gingival inflammation and the presence of periodontitis, suggesting a role for inflamed periodontal tissues as a bacterial gateway into the bloodstream [33].

Despite these limitations, this study was valuable in proposing a structured approach adapted to clinical practice for the evaluation of PODFI in medically compromised patients for whom monitoring of these PODFI must be anticipated for the management of their systemic disease.

These results opened new possibilities for optimizing periodontal assessment protocols, particularly through the integration of standardized measures and appropriate risk criteria into a comprehensive patient-centered approach.

Finally, the results obtained in this study suggested that the proposed periodontal examination and follow-up form could serve as a relevant tool for the diagnosis and infectious monitoring of patients with periodontitis. Its use could enable standardized clinical evaluation and be considered on a national scale, subject to further validation. To validate the different PODFI risk levels, a clinical study examining bacteremia in patients at each level of periodontal infection is necessary.

Periodontal treatment aimed at stabilizing periodontitis and reducing PODFI remains the treatment of choice, even when facing urgent medical milestones. In our practice, patients with this type of clinical history were treated with an initial session comprising an assessment combined with education on hygiene practices and a first full-mouth subgingival debridement performed at the same time, followed by follow-up visits at one-to two-week intervals for a second full-mouth subgingival debridement session. A second study verifying the efficacy of full-mouth treatment in this population of patients with multiple comorbidities could be considered.

## Conclusion

The objective of this study was to assess the severity of PODFI in medically compromised patients using a new periodontal examination and follow-up form, against the backdrop of recent changes in periodontal classifications and the subsequent lack of updated recommendations from the SFCO regarding the management of these infectious foci.

The results highlight a high prevalence of active periodontitis, predominantly at stages III and IV, associated with persistent inflammatory activity. Most patients had a moderate PODFI risk level, while approximately one-third were classified as high risk, underscoring the frequency of periodontal infection foci in patients with high-risk systemic conditions.

Analysis of clinical parameters confirms the central role of biofilm in maintaining periodontal inflammation and in stratifying infectious risk. In patients facing urgent medical milestones, high-risk profiles were primarily associated with significant biofilm accumulation, suggesting that rapid and effective plaque control could help reduce infectious risk prior to certain medical interventions.

Data from the newly developed periodontal assessment allow for the structured evaluation of periodontal infection foci by integrating clinical parameters and potential infectious consequences. Its ability to stratify infection foci could make it a relevant clinical decision-making tool for guiding therapeutic priorities, thereby helping to prevent complications. In daily practice, the integration of such an assessment could improve the management of these patients by facilitating communication among practitioners, optimizing therapeutic decision-making, and allowing for better adaptation of treatment strategies.

However, due to the single-center nature of the study, the small sample size, and the lack of direct bacteremic validation, these results should be interpreted with caution. Prospective multicenter studies appear necessary to confirm the clinical relevance of the proposed risk levels and to validate their association with actual systemic infection risk.

Ultimately, this work highlights the value of updating existing recommendations to incorporate recent advances in periodontology, in order to provide standardized tools adapted to the complexity of clinical situations encountered.

## Declarations

### Ethics approval and consent to participate

The study protocol was submitted to the Data Protection Officer (DPO) of the Lille University Hospital and received a favorable opinion from the institution’s ethics committee (reference: 2025-930-S140). In accordance with French regulations on retrospective research using health data, patients were informed through an information letter available on the Lille University Hospital website and could exercise their right to object to the use of their data.

### Data availability

[To be completed — e.g., The pseudonymized data supporting the findings of this study are available from the corresponding author upon reasonable request, subject to approval by the Lille University Hospital Data Protection Officer.]

### Author contributions

Author contributions are detailed according to the CRediT taxonomy on the title page.

### CRediT

Conceptualization: MB (equal), EG (equal); Methodology: MB (equal), EG (equal); Data curation: CR (equal), SR (equal); Formal analysis: MB; Investigation: CR (lead), SR (lead), MB (support), EG (support); Visualization: MB; Supervision: MB (equal), EG (equal); Writing – Original Draft: CR; Writing – Review & Editing: EG (lead), MB (support)

All authors revised the paper critically for intellectual content and approved the final version. All authors agree to be accountable for the work and to ensure that any questions relating to the accuracy and integrity of the paper are investigated and properly resolved.

